# Mitochondrial DNA copy number is associated with cognitive function, cognitive decline, and dementia: a longitudinal study in UK Biobank

**DOI:** 10.1101/2025.04.11.25324538

**Authors:** Charley Xia, Xinru Su, David C. M. Liewald, Sarah J. Pickett, Gavin Hudson, W. David Hill

## Abstract

Mitochondria are the principal generators of ATP, playing a crucial role in brain health. Mitochondrial DNA copy number (mtDNA-CN) is a proxy for mitochondrial function and has been linked to cognitive function and impairment. However, existing studies are often limited by cross-sectional designs, small sample sizes, or a focus on dementia patients. Using data from 239,070 UK Biobank participants of European ancestry, we examined the longitudinal relationship between mtDNA-CN in blood and cognitive function, as well as its association with cognitive decline, including incident dementia, over a 16-year follow-up. Additionally, we leveraged publicly available genome-wide association study (GWAS) summary statistics to assess the genetic relationship between mtDNA-CN and cognitive function. We found that a higher mtDNA-CN was positively associated with a greater level of cognitive function. Moreover, a higher baseline mtDNA-CN was protective against cognitive decline and all-cause dementia over the 16-year follow-up. However, the effects of mtDNA-CN observed at population-level differed from those observed in individuals experiencing cognitive decline, suggesting heterogeneous influences of mtDNA-CN. Genetic analyses revealed a moderate degree of shared genetic architecture between mtDNA-CN and cognitive function, with overlapping genes involved in pathways associated with known mitochondrial diseases. However, no casual effect of mtDNA-CN was observed on cognitive function. Our findings support mtDNA-CN as a potential biomarker for cognitive health and cognitive ageing. The observed heterogeneity suggests that mtDNA-CN may influence cognition through distinct pathways in normal and cognitively declining individuals. Further research is needed to establish the underlying mechanisms.

## Introduction

Cognitive function is an umbrella term that encompasses multiple correlated cognitive domains including memory, reasoning, processing speed, learning, and problem-solving^1, 2^. Cognitive function is robustly linked with health outcomes, such as cardiovascular disease and stroke, as well as with all as cause mortality, where those with a greater level of cognitive function enjoy better health and a longer life^3^. In the absence of specific conditions that act to reduce cognitive function, cognitive function will decline from middle to older age. This age-related cognitive decline represents the most feared aspect of growing old and can lead to dementia in later life^4, 5^. All-cause dementia affects approximately 2.7% of individuals aged 50–59, with prevalence rising dramatically to 65.9% in those aged 100 and older^6^. By 2050, it is projected that there will be 152.8 million dementia cases globally^7^, underscoring the importance of understanding the aetiology of differences in cognitive function, and in identifying biomarkers for the rate of cognitive decline in older age.

Mitochondrial function as the fundamental biological mechanism of cognitive function has been hypothesised for years^8^ and there are growing evidence of the involvement of mitochondria in neurodegenerative diseases^9–11^. Mitochondria are maternally inherited intracellular organelles that play a fundamental role in cellular energy production through oxidative phosphorylation (OXPHOS)^12,13^. The function of mitochondria also impairs with age, resulting in reduced oxidative capacity and ATP generation, along with increased production of reactive oxygen species (ROS)^14^. As the principal generators of ATP, mitochondrial dysfunction is closely associated with various ageing-related diseases, including chronic respiratory diseases, cancers, and metabolic disorders^15–20^. Since the brain and neurons are amongst the most energy-demanding organ and cell types^21, 22^, mitochondria function is essential for maintaining brain health and its dysfunction is implicated in neurodegenerative diseases including Alzheimer’s disease (AD) and Parkinson’s disease (PD), and Lewy body dementia^23–26^. Although the exact mechanisms remain unclear, disruptions in the PGC-1α-NRF-1/2-TFAM mitochondrial biogenesis pathway, which is closely linked to synaptic activity, are suspected to play a critical role^11^.

Mitochondrial function can be assessed in serval ways, one of which is measuring mitochondrial DNA copy number (mtDNA-CN). mtDNA-CN reflects the quantity of mitochondria in a cell and can be expressed as the amount of mitochondrial DNA relative to nuclear DNA^27^. mtDNA-CN varies upon tissue and cell-type^28, 29^. Previous research has linked blood-derived mtDNA-CN to cognitive function and cognitive impairments^30–33^. However, these studies often limited by cross-sectional designs, small sample sizes, or a narrow focus on dementia patients, whereas the longitudinal effects of mtDNA-CN on individual differences in cognitive function remained largely unexplored.

In this study, we explored the longitudinal phenotypic relationship between mtDNA-CN and cognitive function in 239,070 UK Biobank participants of European ancestry using mtDNA-CN derived from whole-exome sequencing (WES) data from blood samples collected at recruitment and cognitive function assessed using the fluid intelligence test (also called the verbal and numeric reasoning test). This dataset allowed us to investigate the effect of mtDNA-CN on cognitive function at the population level, as well as within sub-populations of individuals experiencing cognitive decline or dementia. Furthermore, by leveraging publicly available genome-wide association study (GWAS) summary statistics, we examined the genetic relationship and overlap between mtDNA-CN and cognitive function.

## Methods

### Ethical Compliance

This UK Biobank project (10279) was approved by the National Research Ethics Service Committee North West-Haydock (REC reference: 11/NW/0382). An electronic signed consent was obtained from all participants.

### Mitochondrial DNA Copy Number

Between 2006 and 2010, 502,655 individuals aged 40–69 years (mean age: 56.5 years) participated in the UK Biobank study^34, 35^ with blood samples collected at the baseline assessment centre visit (**AC1**). Of these, whole-exome sequencing (WES) data were available for 469,913 participants^36^.

Mitochondrial DNA copy number (mtDNA-CN) was derived at a single time point (baseline **AC1**) using WES CRAM files. Note, although no WES probes were originally designed to capture mtDNA, mitochondrial genome can be assembled and mtDNA can be quantified using off-target reads due to the cross-hybridisation of mtDNA with probes that overlap with nuclear-mitochondrial segments (NUMTs)^37–39^. Following the methodology described in Longchamps *et al.*^40^, we summarised read count information for the remaining participants using samtools^41^ and calculated initial mtDNA-CN estimates by regressing the number of reads mapped to the mitochondrial genome against total reads, unknown reads, and decoy reads.

A total of 50,913 participants who had extreme blood counts as defined in Longchamps *et al.*^40^, were not of European ancestry, had mismatched genetic and self-reported sex, or exhibited sex chromosome aneuploidy were excluded. European ancestry was defined as participants whose measure on the first six principal components (PCs), derived using common genotyped nuclear DNA single nucleotide polymorphisms (SNPs), were within 3 standard deviations of the mean for the self-reported European population, and who carried one of the ten common European mitochondrial haplogroups as described in Xia *et al.*^42^.

Next, due to sex-by-age interaction effects^39^, a sex-stratified quality control procedure was performed. Initial mtDNA-CN estimates were regressed on age at baseline, age squared, assessment centre, and mitochondrial haplogroups separately for male and female samples. The standardised residuals from these regressions were obtained, and outliers (residuals greater than 3 standard deviations) were removed. The remaining standardised residuals from both sexes (*N_Male_* = 180,110 and *N_Female_* = 212,389) were then combined and used as the mtDNA-CN phenotype in this study.

### Cognitive Function

Cognitive function data was available for 305,322 participants. It was assessed using the verbal and numerical reasoning test (VNR) consisting of 13 multiple-choice questions covering a range of verbal and numerical problems (UKB Field IDs 20016 and 20191). Participants were given a 2-minute time limit to answer as many questions as possible, and a summed score was calculated based on the total number of correct answers. The VNR test has been shown to have good test-retest reliability (*r* = 0.61) and correlate highly with a general factor extracted from a battery of cognitive tests (*r* = 0.53)^43^.

A total number of 565,965 tests were conducted. These tests were administered on six different occasions: the baseline assessment centre visit (**AC1**), the repeated assessment centre visit (**AC2**), the first imaging centre visit (**AC3**), the repeated imaging centre visit (**AC4**), the first online test (**OL1**), and the repeated online test (**OL2**). Although the online version of the test included an additional 14^th^ question, only three participants answered it correctly within the time limit. As a result, this question was excluded from our analysis to ensure consistency of the test across all six testing occasions.

### Dementia and Parkinson Disease

All-cause dementia (*N* = 10,045) and Parkinson’s disease (*N* = 4,414) cases were identified through self-reports, hospital records, and death records (UKB Field IDs 42018 and 42032). These participants were excluded from the analysis of mtDNA-CN in relation to cognitive function and cognitive decline. However, in the analysis of mtDNA-CN and dementia risk, only dementia cases diagnosed prior to the baseline **AC1** assessment were excluded, while incident dementia cases were retained.

### Phenotypic Analysis

Phenotypic analyses were performed in R v4.2.2. Pearson correlation was computed to evaluate the correlation between cognitive function measured at two different time points using the *cor()* function, whereas Biserial correlation was calculated to assess the correlation between mtDNA-CN and responses to individual test questions using the *polyserial()* function.

Linear regression was employed to estimate the association of mtDNA-CN with cognitive function and cognitive decline using the *lm()* function. The general form of the linear model is as follows:

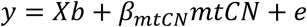

Here, *y* is a vector of one of the followings, cognitive function score, decline in cognitive score, or decline in cognitive score per year, *X* is the design matrix of covariates, *b* is a vector of the corresponding effect sizes for the covariates, *mtCN* is a vector of mtDNA-CN with *β* as the effect of interest, and *e* represents residuals unexplained by the model. Basic covariates included sex, assessment centre, and age at the time of cognitive testing. Where specified in the main text and supplement, additional covariates were fitted in the model, including cognitive function score at baseline, time intervals between tests, major blood composition metrics (red blood cell counts, white blood cell counts, and platelet counts), and smoking status. The null hypothesis (*H*_0_) is *β*_*mtCN*_ = 0 and the alternative hypothesis (*H*_*A*_) is *β*_*mtCN*_ ≠ 0.

Logistic regression was applied to estimate the effects of mtDNA-CN on the odds of exhibiting cognitive decline and incline, and developing dementia using the *glm()* function. Cox proportional hazards model was used to study the difference in dementia risk across age groups using *survival* and *survminer* packages. These models took a similar form as the linear regression model and fitted the same covariates. A flowchart demonstrating the analytic plan and data processing is available in **Supplementary Figure 1**.

### Genetic Analysis

Linkage Disequilibrium Score Regression (LDSC)^44, 45^ was employed to estimate the heritability of, and genetic correlation between, mtDNA-CN and cognitive function. MRlap^46^ was applied to perform two sample Mendelian Randomisation to explore potential causal relationship between mtDNA-CN and cognitive function while accounting for sample overlap, weak instrument bias, and winner’s curse. Functional Mapping and Annotation of Genome-Wide Association Studies (FUMA)^47^ was used to annotate trait-associated loci, identify associated protein coding genes using MAGMA^48^ gene analysis, and detect overrepresented gene sets using hypergeometric tests with all protein-coding genes as the background. To account for multiple testing, false discovery rate (FDR) control was applied on MAGMA gene analysis across 19k genes and within-in family FDR control was performed on overrepresentation analysis for biological process gene sets from MSigDB and GWAS catalog gene-sets separately^49, 50^. Genes and gene-sets with adjusted P-values of lower than 5% were reported. Colocalization analysis was conducted in Coloc^51^ to determine whether the same genetic variant or two independent genetic variants contributed to associations with both mtDNA-CN and cognitive function at a specific locus. SUPERGNOVA^52^ estimated the local genetic covariance between mtDNA-CN and cognitive function across 2,353 independent LD-blocks, as outlined in Zhang *et al*.^52^. Bonferroni correction was applied on SUPERGNOVA results to account for multiple testing and significant threshold was set to *P* < 0.05 / 2,352. 1000 Genome European LD panel served as the reference panel for LDSC, MRlap, FUMA, MAGMA, and SUPERGNOVA analyses.

## Results

### Data Overview

Mitochondrial DNA copy number (mtDNA-CN) and cognitive function were available for 239,070 participants of European ancestry, who have never been diagnosed with dementia or Parkinson disease in the 16-year follow up period. While mtDNA-CN was only available at the first assessment centre visit (**AC1**), these 239,070 participants completed a total of 422,716 cognitive function tests across six time points with an average score of 5.42 (*SD* =1.87) and an average of 1.77 tests per participant. Sample sizes, timeline, and summary of phenotypes are detailed in **Figure 1a** and **Supplementary Table 1**.

**Figure 1.**
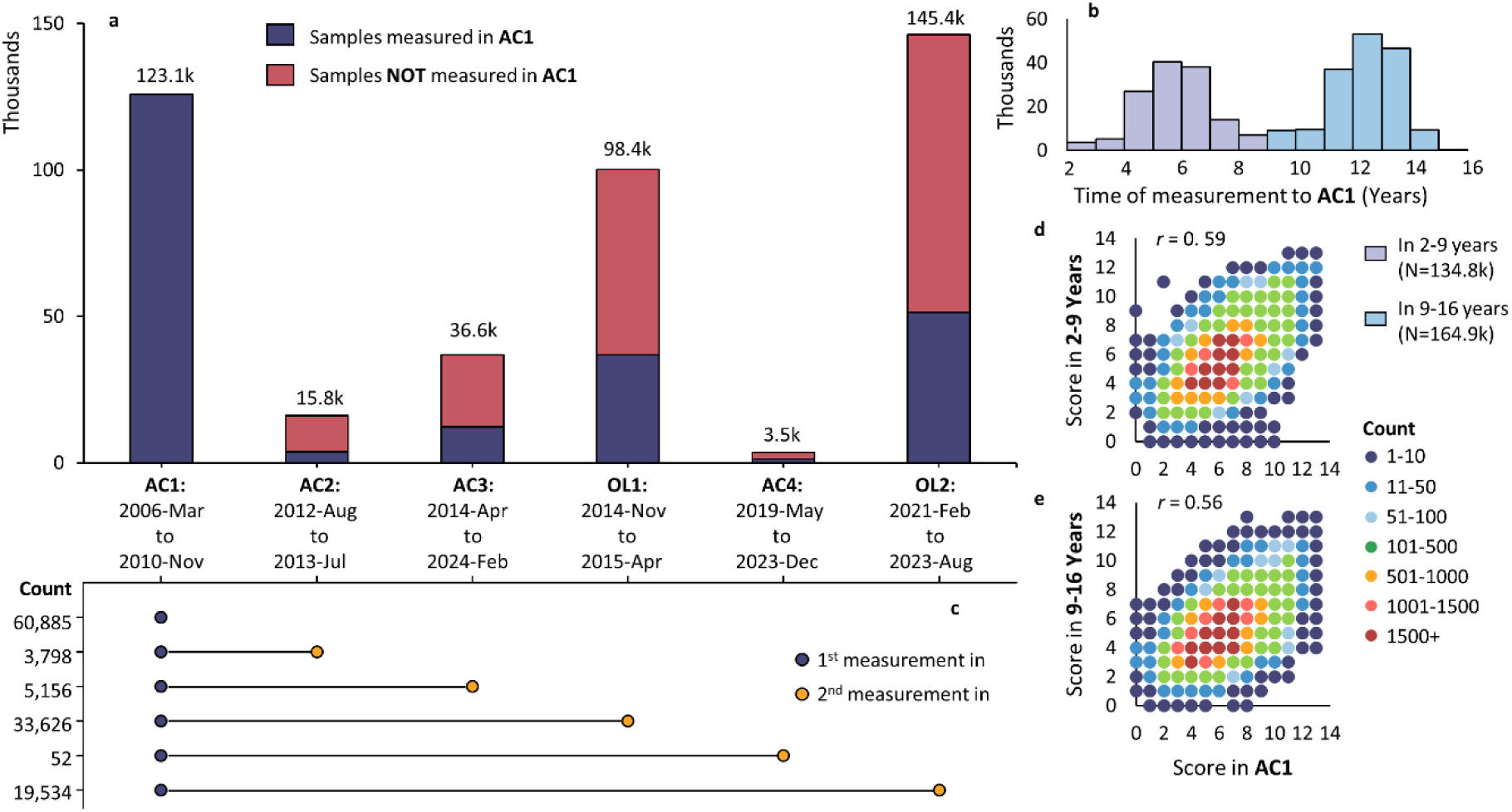
Cognitive Function Sample Size and Correlation. Panel **a**. Sample size (Y-axis) of cognitive function across six time points (X-axis). These six time points are the first (**AC1**) and second (**AC2**) assessment centre visit, the first (**AC3**) and second (**AC4**) imaging centre visit, and the first (**OL1**) and second (**OL2**) online test. These samples have mtDNA-CN measured at **AC1**. Blue bars refer to samples had cognitive function measured at baseline **AC1** whereas red bars do not. Panel **b**. Distribution of time when non-baseline cognitive function was measured, comparing to **AC1**. Panel **c**. Sample size (unique participants) with their first cognitive function measured in **AC1** and a second measurement in any of the rest five time points. Panel **d**. Heatmap and correlation between cognitive function measured in **AC1** and that measured in 2-9 years later. Panel **e**. Heatmap and correlation between cognitive function measured in **AC1** and that measured in 9-16 years later.

Further investigating the time of tests not taken at **AC1**, a bimodal distribution was revealed where 45% of tests (*N_test_* = 134.8k) were performed in 2-9 years after **AC1** (peaking at 5–6 years) and the rest 55% (*N_test_* = 164.9k) were conducted in 9-16 years (peaking at 12–13 years) after **AC1** (**Figure 1b**). These participants were categorised into **2-9 Year** group and **9-16 Year** group accordingly. To avoid having multiple measures per sample, we restricted analysis to the first cognitive measurement at each of the two time periods.

Among the 123,051 participants who had cognitive function measured at baseline (**AC1**), 62,166 (50.5%) completed at least one additional cognitive function test at any of the five subsequent time points (**Figure 1c**). Using these samples, the reliability of cognitive function was assessed by computing the phenotypic correlation between the cognitive function score at **AC1** and those at a subsequent time point. In the **2–9 Year** group (*N* = 41,618), the correlation was 0.593 (95% *CI*: [0.587, 0.599]), consistent with previous findings^43^ and was slightly higher than the correlation of 0.555 (95% *CI*: [0.549, 0.561]) observed in the **9–16 Year** group (*N* = 54,309), with the difference being statistically significant (*z* = 7.68, *P*_diff_ = 8.06 × 10^−15^). Similar results were obtained after adjusting cognitive function for participants’ sex, assessment centre, and age at the time of testing (**Supplementary Table 2**). On average, cognitive function decreased by 0.47 (*SE* = 0.01) in the **2–9 Year** group and by 1.01 (*SE* = 0.01) in the **9–16 Year** group, indicating cognitive ageing.

A flowchart outlining the analytic plan and data processing is available in **Supplementary Figure 1**.

### Positive Association of mtDNA-CN with Cognitive Function at Baseline

We first conducted a cross-sectional analysis to examine the association between mtDNA-CN measured at **AC1** and cognitive function assessed at the same time (*N* = 123.1k). For each test question, a biserial correlation was calculated between mtDNA-CN and score to that question. The analysis revealed that participants with higher mtDNA-CN were more likely to answer questions correctly. These correlations ranged from 0.023 to 0.005 and declined across the 13 questions. Of the 13 correlations, eight were significantly greater than zero at *P* < 0.05. Confidence intervals tended to be wider for both earlier and later questions due to fewer participants answering the earlier questions correctly incorrectly and fewer participants answering the later questions correctly (**Figure 2a** and **Supplementary Figure 2**).

**Figure 2.**
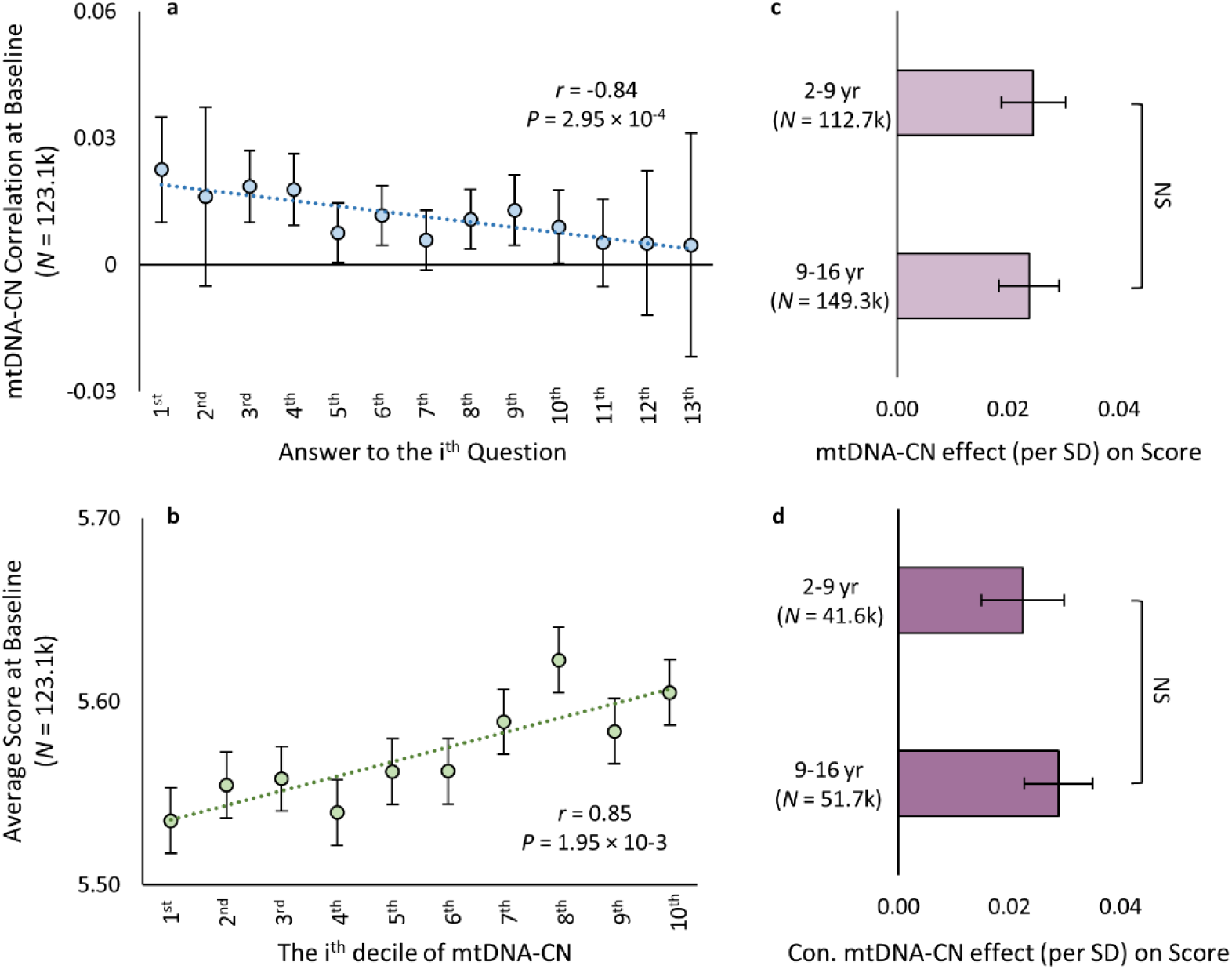
Relationship between mtDNA-CN and Cognitive Function. Panel **a**. Biserial correlation (with 95% confidence interval) between mtDNA-CN and whether answered the i^th^ question correctly in **AC1**. Panel **b**. Average cognitive function (with standard error) across mtDNA-CN deciles. Panels **c-d**. Effects (with standard error) of mtDNA-CN on cognitive function estimated in all participants and 62.2k participants whose first cognitive function was measured in **AC1**, respectively. Y-axis: time when the cognitive test was taken. X-axis: mtDNA-CN effect size per SD. ‘Con.’ refers to the conditioned mtDNA-CN effects where baseline **AC1** cognitive function and time interval between tests were adjusted in the model. ‘NS’ means the between group difference is not significant at *P* < 0.05. In all panels where applicable, dotted line is the fitted line for the plot, *r* is the Pearson correlation between X and Y, and *P* is the p-value of the correlation, and mtDNA-CN decile is ranked from the lowest (1^st^) to the highest (10^th^).

Participants were then grouped into deciles based on their mtDNA-CN and the average score was compared across these groups. A clear positive linear trend was observed between mtDNA-CN and cognitive function, where individuals in the lowest mtDNA-CN decile achieved a score of 5.54 (*SE* = 0.02), those in the highest mtDNA-CN decile had a score of 5.61 (*SE* = 0.02), and per decile increase in mtDNA-CN was associated with an increase of 0.009 (*SE* = 0.002, *P* = 1.95 × 10^−3^) (**Figure 2b**).

To further quantify this association, we performed linear regression analysis. Results showed that a one standard deviation increase in mtDNA-CN was associated with an increase of 0.026 (*SE* = 0.006, *P* = 6.32 × 10^−6^) scores in cognitive function. In this model, participants’ sex, assessment centre, and age at the time of testing were adjusted.

Given that blood composition and smoking are known factors associated with mtDNA-CN^53, 54^, we repeated the linear association by including these variables as additional covariates. The association between mtDNA-CN and cognitive function remained significant after these adjustments with slight attenuation in the effect size (**Supplementary Table 3**), indicating that the observed mtDNA-CN effects on cognitive function were not solely mediated through these factors.

### mtDNA-CN at Baseline Predicts Future Cognitive Function and Cognitive Change

We next performed a longitudinal analysis to investigate the association between mtDNA-CN measured at baseline **AC1** and cognitive function assessed at subsequent time points. Using the same linear regression model, we discovered that a one standard deviation increase in mtDNA-CN at **AC1** was associated with an increase of 0.025 (SE = 0.005, *P* = 6.76 × 10^−6^, *N* = 112.7k) scores in cognitive function in **2–9 Year** group and an increase of 0.024 (SE = 0.004, *P* = 3.37 × 10^−8^, *N* = 149.3k) scores in **9–16 Year** (**Figure 2c**).

Since cognitive function measured at later time points is highly correlated with the baseline measurement (**Figures 1d-e**), we performed additional analyses to ensure the observed effects of mtDNA-CN on subsequent cognitive function were not solely driven by baseline ability. Using the 62.2k participants with at least two cognitive function assessments (one at baseline **AC1** and one at a subsequent time point), we re-examined the association between mtDNA-CN and the second cognitive function measurement while conditioning on the baseline measure and the time interval between two tests. The conditioned effect of mtDNA-CN remained significant, with positive effects of 0.022 (*SE* = 0.007, *P* = 2.81 × 10^−3^, *N* = 41.6k) scores in the **2–9 Year** group, 0.029 (*SE* = 0.006, *P* = 3.15 × 10^−6^, *N* = 51.7k) scores in the **9–16 Year** group, and mtDNA-CN explained up to 0.03% of the model variance (**Figure 2d** and **Supplementary Table 4**). These results indicate that the mtDNA-CN measured at baseline **AC1** could predict the change in cognitive function in the follow-up period.

We further adjusted the regression models for blood composition and smoking. The effects remained significant (**Supplementary Tables 3-4**).

### Higher mtDNA-CN Protects against Cognitive Decline and Dementia but is Not Associated with Practice Effects

To further investigate the relationship between mtDNA-CN and cognitive change, we classified the 62.2k participants with at least two cognitive assessments into a ‘decline’ group (*N* = 19.7k) and an ‘incline’ group (*N* = 3.9k), defines by score changes exceeding ±1 SD between baseline **AC1** and follow-up. For participants with two subsequential measures (*N* = 31.2k), the change had to be continuous (see **Supplementary Figure 3** for illustration).

Participants with higher mtDNA-CN were less likely to experience cognitive decline. Specifically, 30.8% of participants in the highest decile exhibited cognitive decline, compared to 32.6% in the lowest decile, with a 0.15% decrease in prevalence per decile (*SE* = 0.05%, *P* = 0.03, **Figure 3a**). Logistic regression showed a 3.3% reduction in the odds of experiencing cognitive decline per 1 SD increase in mtDNA-CN (*OR* = 0.967, 95% *CI*: [0.948, 0.986], *P* = 8.72 × 10⁻⁴), and an 8.6% reduction for those in the highest decile versus the lowest decile (*OR* = 0.914, 95% *CI*: [0.839, 0.996], *P* = 0.04), over a 16-year follow-up period (**Figure 3b**). On the contrary, no significant effects were found for the ‘incline’ group (**Figures 3a-b**).

**Figure 3.**
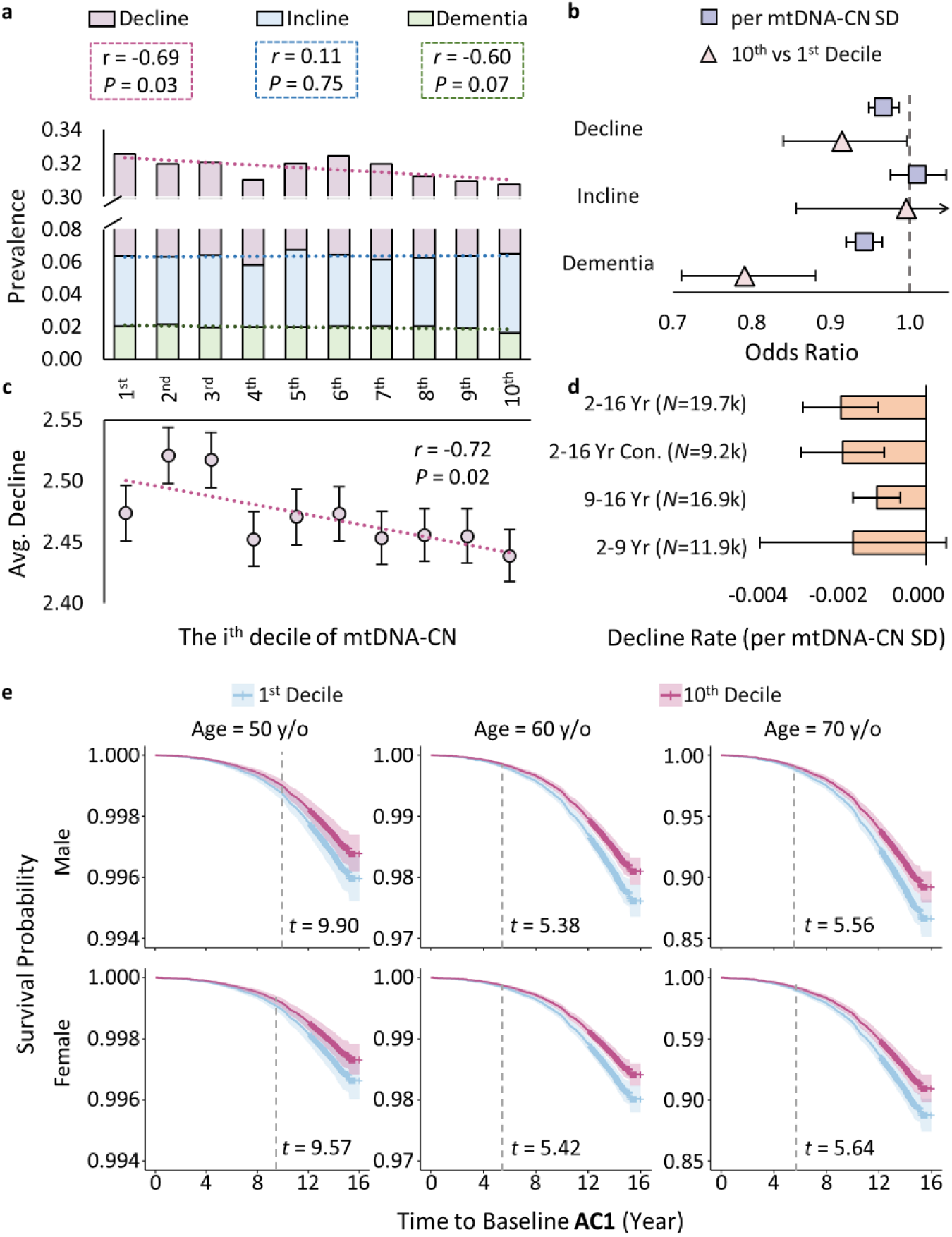
Relationship between mtDNA-CN and Cognitive Decline and Dementia. Panel **a**. Prevalence of ‘decline’, ‘incline’, and dementia (Y-axis) across mtDNA-CN deciles (X-axis). Panel **b**. Logistic regression results showing the odds ratios (with standard error) for mtDNA-CN predicting risk of ‘decline’, ‘incline’, and dementia in the entire16-year follow-up. Panel **c**. Average cognitive decline (with standar error) across mtDNA-CN deciles in participants identified as having ‘decline’. Panel **d**. Effects (with standard error) of mtDNA-CN on cognitive decline rate across different ‘decline’ groups and time periods. ‘2-16 Yr’, ‘9-16 Yr’, and ‘2-9 Yr’ represent estimates from the respective time windows. ‘2-16 Yr Con.’ refers to participants with continuous declines from baseline **AC1** to 2-9 year and from 2-9 year to 9-16 year. Panel **e**, Survival probability (with 95% confidence interval) of dementia-free where dotted line indicates the earliest time point where the differences between the 1^st^ and 10^th^ mtDNA-CN deciles significantly differ at *P* < 0.05. In panels **a**-**c**, dotted line is the fitted line for the plot, *r* is the Pearson correlation between X and Y, and *P* is the p-value of the correlation. In all panels where applicable, mtDNA-CN decile range from the lowest (1^st^) to the highest (10^th^).

Participants with higher mtDNA-CN also had a shallower rate of decline in cognitive function. Within the ‘decline’ group (*N* = 19.7k), a negative correlation was observed between mtDNA-CN decile and the magnitude of cognitive decline (*r* = −0.72, 95% *CI*: [0.16 – 0.93], *P* = 0.02, **Figure 3c**). Linear regression showed that 1 SD increase in mtDNA-CN was associated with a 0.015-score milder decline (SE = 0.006, *P* = 0.017) and 0.002 less decline per year (SE = 0.006, *P* = 0.017) over the entire 16-year follow-up. Similar levels of protective effects were observed in different time windows and in a more conservative ‘decline’ subset consisting of only participants with continuous decline from baseline **AC1** to 2-9 year and from 2-9 year to 9-16 year (**Figure 3d** and **Supplementary Figure 4**). Note, as the magnitude of cognitive decline increases with longer intervals between assessments, combing measurements taken at varying time periods can obscure the detection of mtDNA-CN effects. To reduce this variability, for participants with two subsequential measures (*N* = 31.2k), we took the measurements best aligned with the time period for the analysis (**Supplementary Table 5** and **Supplementary Figures 3 & 5**).

The protective effects of mtDNA-CN on cognitive decline also extend to pathological cases. We recovered 153,089 European participants with mtDNA-CN data that were excluded in the original analysis, including 7,721 incident dementia. In this expanded cohort (*N* = 392,159), mtDNA-CN decile was negatively correlated with the dementia prevalence (*r* = −0.60, 95% *CI*: [-0.89, 0.05], *P* = 0.02, **Figure 3a**). Over a 16-year follow-up period, each 1SD increase in mtDNA-CN was associated with a 5.7% reduction in the odds of developing dementia (*OR* = 0.943, 95% *CI*: [0.920, 0.966], *P* = 1.60 × 10^−6^), and a 20.9% reduction in the odds was observed for those in the highest decile versus the lowest decile (*OR* = 0.791, 95% *CI*: [0.711, 0.880], *P* = 1.78 × 10^−5^, **Figure 3b**). For participants aged between 50-70 y/o, the risks of developing dementia between the highest and lowest decile became significant 5-10 years after baseline **AC1** (**Figure 3e**).

A sensitivity analysis indicated that the effects of mtDNA-CN on cognitive decline and dementia risk remained consistent following adjustment for blood composition and smoking, (**Supplementary Tables 6-7**).

### Positive Genetic Correlation while No Casual Effects between Two Traits

As both mtDNA-CN and cognitive function are heritable traits^54, 55^, we conducted a genetic study to explore potential mechanism underlying the observed phenotypic associations between them. For this purpose, we used the summary statistics from the Genome-Wide Association Study (GWAS) available: Lee et al.^55^ (*N* = 257.8) on cognitive function and Gupta et al.^54^ (*N* = 274.8k) on blood-derived mtDNA-CN. Both GWAS primarily involved participants from the UK Biobank.

We first used Linkage Disequilibrium Score Regression (LDSC)^44, 45^ to estimate the heritability of cognitive function and mtDNA-CN and assess the genetic correlation between the them. The LDSC heritability was 0.192 ± 0.006 (*P* = 1.47 × 10^−210^) for cognitive function and 0.047 ± 0.004 (*P* = 3.53 × 10^−30^) for mtDNA-CN. The genetic correlation between the two traits was estimated at 0.091 ± 0.027 (*P* = 7.23 × 10^−4^), providing evidence of a small but significant genetic overlap between cognitive function and mtDNA-CN (**Figure 4a**).

**Figure 4.**
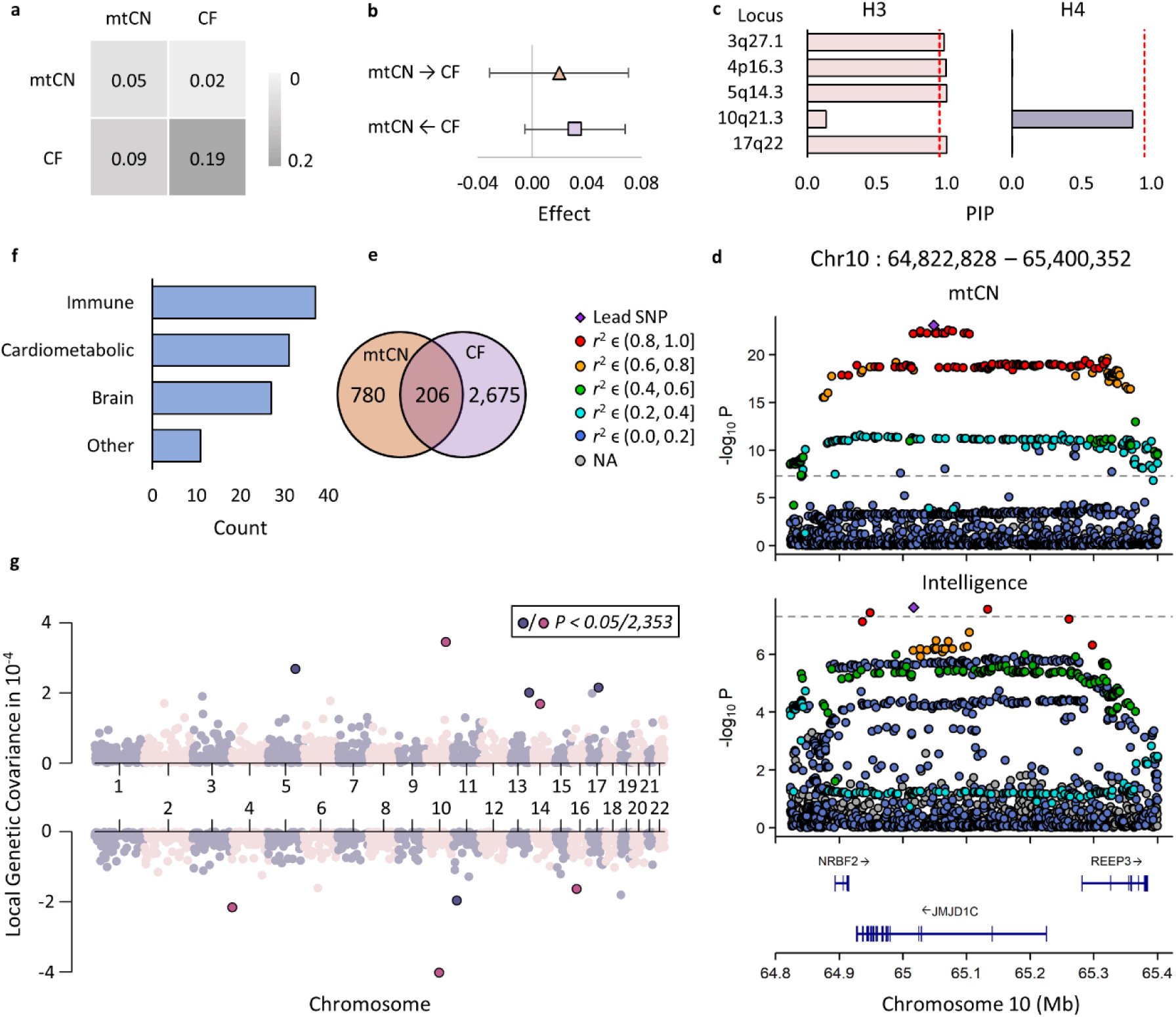
Genetic Overlay between mtDNA-CN and Cognitive Function. Panel **a**, correlation between and heritability of mtDNA-CN and cognitive function (CF). Diagonal: heritability. Lower-diagonal: genetic correlation. Upper-diagonal: phenotypic correlation at **AC1**. Panel **b**, MRlap results with standard error. mtDNA-CN→CF: effect of mtDNA-CN on cognitive function. mtDNA-CN←CF: effect of cognitive function on mtDNA-CN. Panel **c**, colocalization hypothesis testing results. *H*_3_: both traits are associated, but have different single causal variants (left). *H*_4_: both traits are associated and share the same single causal variant (right). X-axis: posterior inclusion probability (PIP). Red dotted line: suggestive significant threshold of 0.95. Panel **d**, Locus zoom plots for mtDNA-CN (top) and cognitive function (bottom) on genomic overlap region chr10:64,822,828–65,400,352. Purple diamonds represent the lead SNPs with the lowest association p-value. Dots represent other SNPs in the same region and are coloured based on their LD with the lead SNP. Grey dots refer to SNPs with no LD information in the 1000 Genome European reference panel. Panels **e**, Venn diagram shows the overlap between mtDNA-CN-associated genes and cognitive function -associated genes. Panels **f**, Count of overrepresented GWAS catalog gene-sets in four domains. Panels **g**, local genetic covariance between mtDNA-CN and cognitive function across 2,353 independent LD-blocks in the genome. Each dots represent a LD-block and is coloured based on odd or even chromosome. X-axis, chromosome number and relative position on the chromosome. Y-axis, estimate of local genetic covariance in 10^−4^. Darker colour and black-border are used for LD-block that explains significant amount of local genetic covariance at *P* < 0.05/2,353.

Subsequently, we conducted a two-sample Mendelian Randomisation (MR) analysis, treating mtDNA-CN as the exposure and cognitive function as the outcome, and vice versa, to explore potential causal effects (**Figure 4b**). Since both GWAS included UK Biobank samples, we accounted for sample overlap using Mrlap^46^. MR results showed no causal effects of mtDNA-CN on cognitive function (0.020 ± 0.025, *P* = 0.44) nor casual effects of cognitive function on mtDNA-CN (0.031 ± 0.018, *P* = 0.09).

### Few Shared Loci, Moderate Number of Shared Genes and Bidirectional Local Genetic Covariance between mtDNA-CN and Cognitive function

To further explore the relationship between mtDNA-CN and cognitive function, we investigated genetic overlap at the locus, gene, and regional levels.

First, we annotated the GWAS summary statistics for both traits using FUMA^47^ to identify trait-associated loci and identified 79 loci associated with mtDNA-CN and 183 loci associated with cognitive function (**Supplementary Tables 8–9**). Among these, ten loci physically overlapped (five from each trait), forming five shared genomic regions: chr3:18,4061,387-184,093,040, chr4:696,848–730,476, chr5:87,936,379–88,416,354, chr10:64,822,828–65,400,352, and chr17:56,325,771–57,229,716. For each overlapping region, we performed colocalization analysis using coloc^51^ to determine whether the GWAS signals for the two traits were driven by the same underlying genetic variant or by independent variants. The results showed that for regions on chromosomes 3, 4, 5, and 17, there was a posterior probability exceeding 0.95 that independent genetic variants contributed to the associations with the two traits. In contrast, for the region on chromosome 10, there was some evidence that a shared genetic variant was responsible for the observed associations (**Figures 4c-d**).

Next, we used MAGMA^48^ to identify trait-associated genes at FDR of 5%. We identified 780 genes associated with mtDNA-CN and 2,675 genes associated with intelligence, with 206 genes shared between the two traits (**Figure 4e** and **Supplementary Tables 10–12**). To explore the functions of these shared genes, we performed hypergeometric tests to determine whether they were enriched in predefined gene sets from MsigDB and the GWAS Catalog^49, 50^ at FDR of 5%. The results showed significant enrichment of these genes in the biological pathway of homophilic cell adhesion via plasma membrane adhesion molecules (*P_adj_* = 6.89 × 10^−3^) and in 106 published GWAS (**Figure 4f**). These 106 GWAS include 37 immune-related GWAS, such as blood cell traits, asthma, and inflammatory bowel disease; 27 brain-related GWAS, such as intelligence, brain morphology, personality, and mental health; 31 cardiometabolic GWAS, such as BMI, diabetes, coronary artery disease, and blood pressure; and 11 uncategorised GWAS including cancer and longevity (**Supplementary Table 13**).

Lastly, we estimated the local genetic covariance between mtDNA-CN and cognitive function across 2,353 pre-defined independent LD-blocks in the genome using SUPERGNOVA^52^ (**Figure 4g** and **Supplementary Table 14**). Nine genomic regions exhibited significant genetic covariance at *P* < 0.05/2,353 level. In contrast to the overall positive genetic correlation observed in LDSC, the local genetic covariance between the two traits was bidirectional. Specifically, four of the nine regions showed negative covariance. Notably, the overlapping genetic locus on chromosome 10 that likely harboured a shared underlying variant (**Figure 4d**) was located in a region, chr10:64,263,402– 65,469,133, that explained the largest amount of negative covariance. In that shared locus, both lead SNPs of rs7080386 for mtDNA-CN and rs7896910 for cognitive function are intron variants of *JMJD1C*, a gene involved in a genetic neurodevelopmental disorder named Rett Syndrome^56^. Rett Syndrome patients demonstrated different levels of cognitive impairments^57^ and an increased amount of mtDNA-CN^58^, explaining the observed negative local genetic covariance.

## Discussion

Maintaining cognitive ability is essential for maintaining independence and quality of life in older age. Cognitive decline significantly impacts well-being, health, and adds an increasing financial burden on healthcare systems. Previous studies have shown that patients with Alzheimer’s disease harbour lower mitochondrial DNA copy number (mtDNA-CN) compared to non-affected individuals^30, 59^ and mtDNA-CN may have a causal effect on dementia^60^. In this study, we conducted comprehensive analyses to explore the relationship between mtDNA-CN and cognitive function in a non-clinical population, providing new insights into their phenotypic associations, genetic underpinnings, and the potential mechanisms that influence cognitive function and its decline.

Our phenotypic analysis revealed that mtDNA-CN recorded at baseline is positively associated with cognitive function and cognitive change (**Figure 2**). Specifically, a one standard deviation increase in mtDNA-CN at baseline was associated with a 0.02 - 0.03 scores increase in cognitive function measured 2–16 years later, with and without adjusting for the baseline level of cognitive function. This finding complemented prior research, which identified a link between baseline mtDNA-CN and general cognitive function over a 5–20-year period without accounting for baseline cognition^31^. Furthermore, we found that higher baseline mtDNA-CN levels protect against cognitive decline and all-cause dementia (**Figure 3**). Specifically, comparing to the lowest mtDNA-CN decile, being in the highest mtDNA-CN decile at baseline reduces the odds of having cognitive decline by 8.6% and having all-cause dementia by 20.9% in 16-year follow-up period. Notably, the observed association of mtDNA-CN with change in cognitive function only presented in cognitive decline group, suggesting potential heterogeneous effects of mtDNA-CN that operate differently in individuals with normal cognitive function versus those experiencing cognitive decline, highlighting the potential role of mtDNA-CN in neurodegenerative process and as a biomarker for long-term cognitive decline and neurological health.

Our genetic analyses revealed the shared genetic architecture between mtDNA-CN and cognitive function at various levels, including loci, genes, local regions, and the genome as a whole (**Figure 4**). The estimated genetic correlation between the two traits was small but significant (0.091 ± 0.027, *P* = 7.23 × 10^−4^), a finding that is attributable to two factors. First, there is limited evidence for shared genetic variants directly influencing both traits as indicated by colocalization results. Second, the bidirectional local genetic covariance observed suggests the presence of two etiologically distinct underlying mechanisms driving genetic associations in opposite directions. Negative local genetic covariance may also explain why in previous studies an increased mtDNA-CN was found associated with other diseases, such as some cancers, where compensatory effects present^61^. Despite having little shared genetic variants and the absence of a consistent direction of SNP effects as indicated by the bidirectional local genetic covariance, we identified more than 200 genes associated with both traits, implicating shared biological pathways. These pathways are not only vital for cognitive function but also play significant roles in brain morphology, mental health, cardiovascular diseases, immune system disorders, and metabolic syndromes—domains consistently linked to mitochondrial function (**Supplementary Table 13**). These common genes highlight the importance of further investigating the genetics of mtDNA-CN and its broader connections to other traits and diseases, extending beyond cognitive function, and bridges the gap from mitochondrial dysfunction -> genetics -> mtDNA-CN -> diseases.

Integrating previous discoveries with our findings, we hypothesise a potential mechanism linking mtDNA-CN to cognitive function and cognitive decline. Mitochondrial dynamics respond to energy demand and nutrition supply^62^. Consequently, mtDNA-CN, unlike other heritable traits such as height that are relatively stable across adult-lifetime, is subject to short term change. For example, studies have shown that weeks of exercise can lead to significant changes in mtDNA-CN in human, blood, brain and muscle^63–66^. When mitochondrial function is impaired, mtDNA-CN can change more rapidly than the time required for cognitive decline or late-onset dementia to manifest. This can disrupt the short-term association between mtDNA-CN and cognitive function while establishing a longer-term link between mtDNA-CN and cognitive decline or dementia incidence in the future. Although all participants appeared cognitively healthy at baseline recruitment, the cohort included both healthy and subclinical individuals, with time-dependent events already influencing baseline mtDNA-CN measures in blood, leading to the heterogeneous phenotypic effects of mtDNA-CN observed in our study. It also provides a rationale for the lack of causal effects of mtDNA-CN on cognitive function detected in our Mendelian randomisation (MR) analysis, consistent with previous publication^31^. Future studies investigating the effects of mtDNA-CN on cognitively healthy and unhealthy participants separately may further elucidate this working theory.

Our study has a several limitations. First, like many studies, we utilised blood-derived mtDNA-CN due to its accessibility. However, mtDNA-CN exhibits tissue and cell-type specificity^28, 29^. As such, mtDNA-CN in blood may fail to capture some aspects of mtDNA-CN in brain and using brain-derived mtDNA-CN could potentially reveal stronger or different associations with cognitive function^30^. Nonetheless, the observed association between blood-derived mtDNA-CN and cognitive function indicates that our blood-based marker could capture the mitochondrial effects relevant to brain tissue.

Second, while our dataset includes multiple measurements of cognitive function over different time points, mtDNA-CN was measured only at baseline recruitment. Although mtDNA-CN is predictive of future cognitive decline, having repeated mtDNA-CN measurements during follow-up periods could provide deeper insights into its long-term effects on future cognitive decline and dementia incidence, further clarifying our proposed theory. In addition, the participants of UK Biobank were of a broad age range at baseline when undergoing cognitive assessment and the interval between cognitive tests also varies between participants. As consequence, this age and interval heterogeneity may serve to occlude the association between cognitive function/cognitive decline and mtDNA-CN. Future work utilising cohorts with a narrower age range, and including cognitive testing at uniform time points, would be valuable in gauging the magnitude of the associations identified here.

Third, our study found a positive association between mtDNA-CN and cognitive function at the population level. However, MR analysis failed to find evidence to support a causal role for mtDNA-CN in this association. Given that mtDNA-CN also influences cognitive decline and that the UK Biobank cohort consists of mainly middle-aged participants^67^ and inevitably included individuals with subclinical conditions, studying a younger and healthier cohort may provide a clearer understanding of the relationship between mtDNA-CN and cognitive function in general population. This could also help disentangle the heterogeneous mtDNA-CN effects observed in our study, further clarifying the proposed mechanism.

Fourth, blood cell-type composition is a significant confounder for blood-derived mtDNA-CN due to its derivation method and necessitates careful adjustment^40, 54^. However, mtDNA-CN is also linked to inflammation^68, 69^, which affects blood cell-type composition and itself is a risk factor for cognitive decline and dementia^70^. As a result, adjusting for blood cell-type composition could potentially obscure true associations between mtDNA-CN and cognitive function if the relationship is primarily mediated through inflammatory pathways. Although our associations remained significant after such adjustments, previous studies have reported that accounting for blood cell-type composition eliminated associations between mtDNA-CN and conditions such as Parkinson’s disease, metabolic traits, and cardiovascular diseases^54, 71, 72^. To better understand the relationship of mtDNA-CN with cognitive function and other diseases, future research should focus on using more refined methods for measuring mtDNA-CN that are less influenced by blood cell-type composition or measuring mtDNA-CN in appropriate tissues.

Future work in large samples where brain tissue is available will be valuable for replicating and extending our findings. However, our study illustrates that mtDNA-CN derived using blood likely influences cognitive function in healthy adults. Furthermore, blood-derived mtDNA-CN can be used as a predictor of future levels of cognitive decline and dementia, highlighting its potential role as a biomarker for disease prediction and prevention.

## Data Availability

UK Biobank data used in this study are available via the UK Biobank data access process (see http://www.ukbiobank.ac.uk/register-apply/). Cognitive function GWAS summary statistics are available at https://thessgac.com/ whereas mtCN GWAS summary statistics are available on GWAS catalog at https://www.ebi.ac.uk/gwas. MRlap is available at https://github.com/n-mounier/MRlap. Coloc is available at https://chr1swallace.github.io/coloc. FUMA is a web-based tool available at https://fuma.ctglab.nl. Supergnova is available at https://github.com/qlu-lab/SUPERGNOVA. LDSC and 1000 Genome European reference panel are available at https://github.com/bulik/ldsc.

https://thessgac.com/

https://www.ebi.ac.uk/gwas

https://github.com/n-mounier/MRlap

https://chr1swallace.github.io/colo

https://fuma.ctglab.nl

https://github.com/qlu-lab/SUPERGNOVA

https://github.com/bulik/ldsc

## Data and Code Availability

UK Biobank data used in this study are available via the UK Biobank data access process (see http://www.ukbiobank.ac.uk/register-apply/). Cognitive function GWAS summary statistics are available at https://thessgac.com/ whereas mtDNA-CN GWAS summary statistics are available on GWAS catalog at https://www.ebi.ac.uk/gwas. Mrlap is available at https://github.com/n-mounier/Mrlap. Coloc is available at https://chr1swallace.github.io/coloc. FUMA is a web-based tool available at https://fuma.ctglab.nl. Supergnova is available at https://github.com/qlu-lab/SUPERGNOVA. LDSC and 1000 Genome European reference panel are available at https://github.com/bulik/ldsc.

## Acknowledgements

C.X. and W.D.H. are supported by a Career Development Award from the Medical Research Council (MRC) [MR/T030852/1] for the project titled “From genetic sequence to phenotypic consequence: genetic and environmental links between cognitive ability, socioeconomic position, and health”. GH receives support from Parkinson’s UK (G-2003), the Michael J Fox Foundation (MJFF-007574 and MJFF-007690) and the Wellcome Centre for Mitochondrial Research (203105/Z/16/Z). S.J.P is supported by a Wellcome Trust Career Re-entry Fellowship (204709/Z/16/Z), the Wellcome Centre for Mitochondrial Research (203105/Z/16/Z) and also receives support from a L’Oreal UNESCO FWIS Award. For the purpose of open access, the author has applied a ‘Creative Commons Attribution (CC BY) licence to any Author Accepted Manuscript version arising from this submission.

