## Supplementary Figures and Tables for "Mitochondrial DNA copy number is associated with cognitive function, cognitive decline, and dementia: a longitudinal study in UK Biobank": SP Figures for NA.docx

### Supplementary Figure 1. Flowchart for data prepration and analytic plan of phenotypic association analysis.

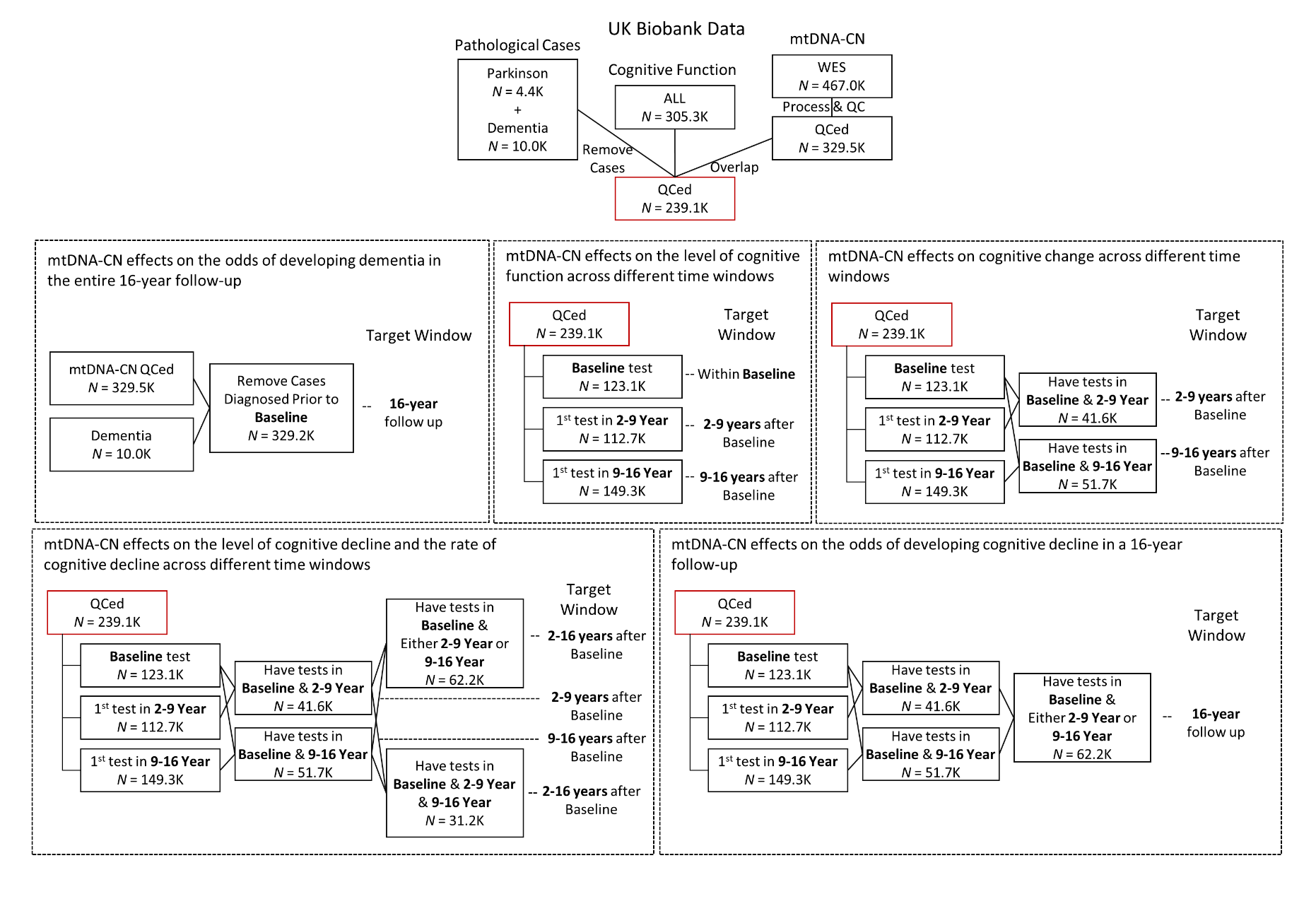

This flowchart shows how we prepared the data and which data sets were used for what analysis.

### Supplementary Figure 2. Relationship between percentage of correct/wrong answers and standard errors of biserial correlation across 13 cognitive function tests.

SE

| Percentage of correct/wrong answers | All |
| --- | --- |
| 4.40093% | 0.00636145 |
| 1.04341% | 0.0107635 |
| 15.8864% | 0.00433132 |
| 16.1654% | 0.00430399 |
| 47.6243% | 0.00357448 |
| 40.5151% | 0.00361378 |
| 36.9446% | 0.003651 |
| 41.8597% | 0.003596 |
| 16.7169% | 0.00422318 |
| 14.2629% | 0.00438956 |
| 7.49303% | 0.00528063 |
| 1.67439% | 0.00872547 |
| 0.533229% | 0.0134609 |

| Percentage of correct/wrong answers | %Correct |
| --- | --- |
| 47.6243% | 0.00357448 |
| 41.8597% | 0.003596 |
| 16.7169% | 0.00422318 |
| 14.2629% | 0.00438956 |
| 7.49303% | 0.00528063 |
| 1.67439% | 0.00872547 |
| 0.533229% | 0.0134609 |

| Percentage of correct/wrong answers | %Wrong |
| --- | --- |
| 4.40093% | 0.00636145 |
| 1.04341% | 0.0107635 |
| 15.8864% | 0.00433132 |
| 16.1654% | 0.00430399 |
| 40.5151% | 0.00361378 |
| 36.9446% | 0.003651 |

The figue shows that standard error goes up when fewer particiapnts answer questions correctly or incorrectly. Y-axis: standard error. X-axis: percentage of correct/wrong answers. When less than 50% of the participants answer the question correctly, percentage of correct answer is used for X-axis, otherwise percentage of wrong answer is used here. Data label: the order of the 13 questions. Dotted line: Fitted line. Rectangle box: equation and R-squared of the fitted line.

### Supplementary Figure 3. Illustration of how to define ‘decline’ and ‘incline’ and how to calculate cognitve change and time interval for different subgroups.

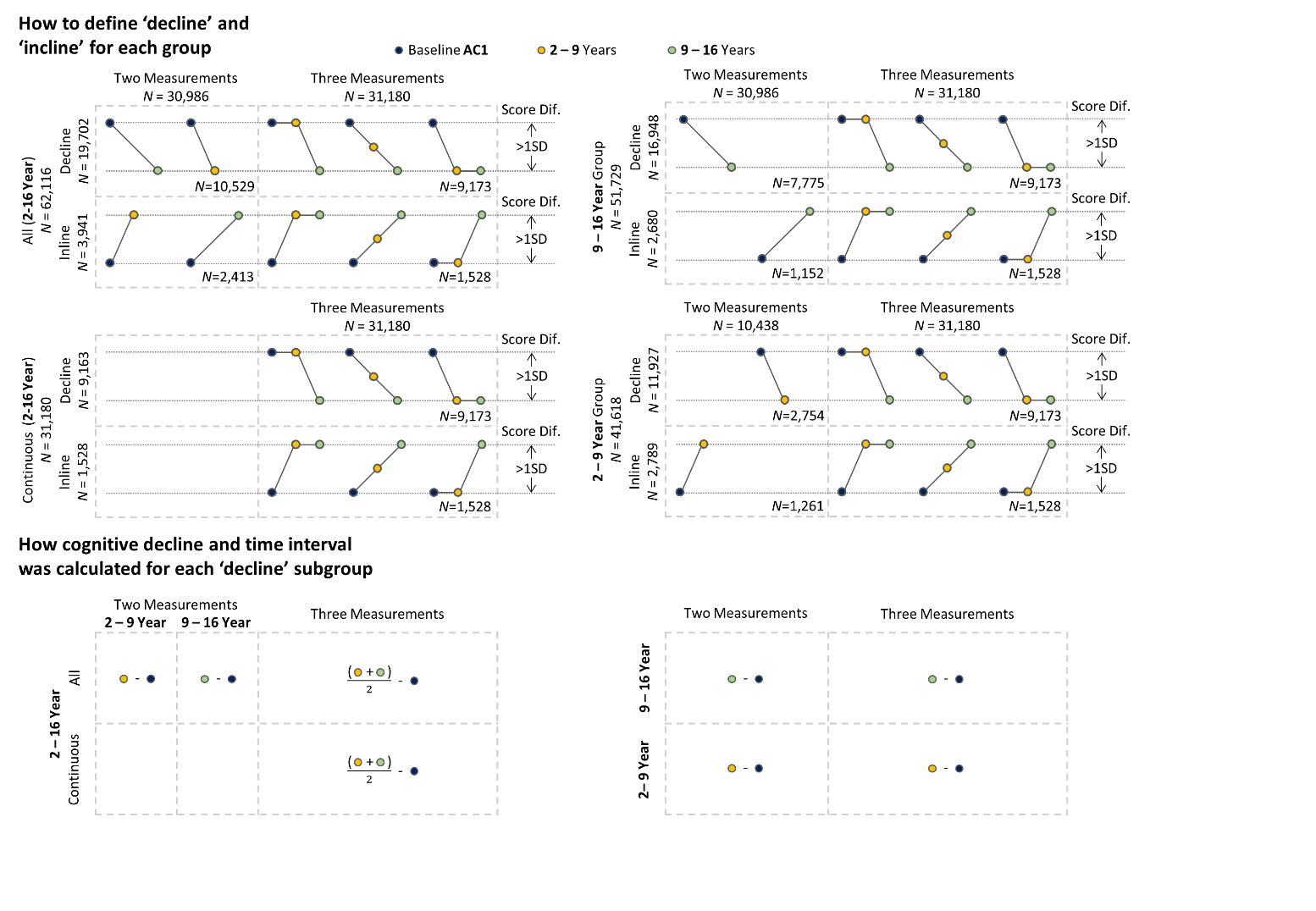

For participants with two cognitive assessments, ‘decline’ or ’incline’ was defined a change from baseline exceeding ±1 SD in test scores (with 1 SD at baseline equal to 1.96). For those with three assessments, the change was defined based on the difference between the last and baseline scores exceeding ±1 SD, with the intermediate score falling between the first and last measurements. In assessing the effects of mtDNA-CN on cognitive decline, the cognitive test score used was matched to the appropriate follow-up interval. Specifically, for 31,180 participants with three measurements, the measurement taken 2–9 years after baseline was used for studying the effects within 2-9 years of follow-up, the measurement taken 9–16 years after baseline was used for studying the effects within 9–16 years of follow-up, and the average score of and the mean time interval between the 2–9 year and 9–16 year assessments were used when analyses covering the entire 16-year follow-up period.

### Supplementary Figure 4. Effects of mtDNA-CN on cognitive decline.

| Subgroup based on time after baseline | Cognitive Decline |
| --- | --- |
| 2-9 Year | -0.0131202 |
| 9-16 Year | -0.0143707 |
| 2-16 Year (Continuous) | -0.0176446 |
| 2-16 Year | -0.015394 |

Figure shows that per SD increase in mtDNA-CN is associated with a milder cognitive decline and the effects are consistent across different follow-up periods.

### Supplementary Figure 5. Distribution of time interval for participants identified as ‘decline’ in each follow-up group using different measurements.

2 – 16 Yr (Continuous)

2 – 16 Year

9 – 16 Year

2 – 9 Year

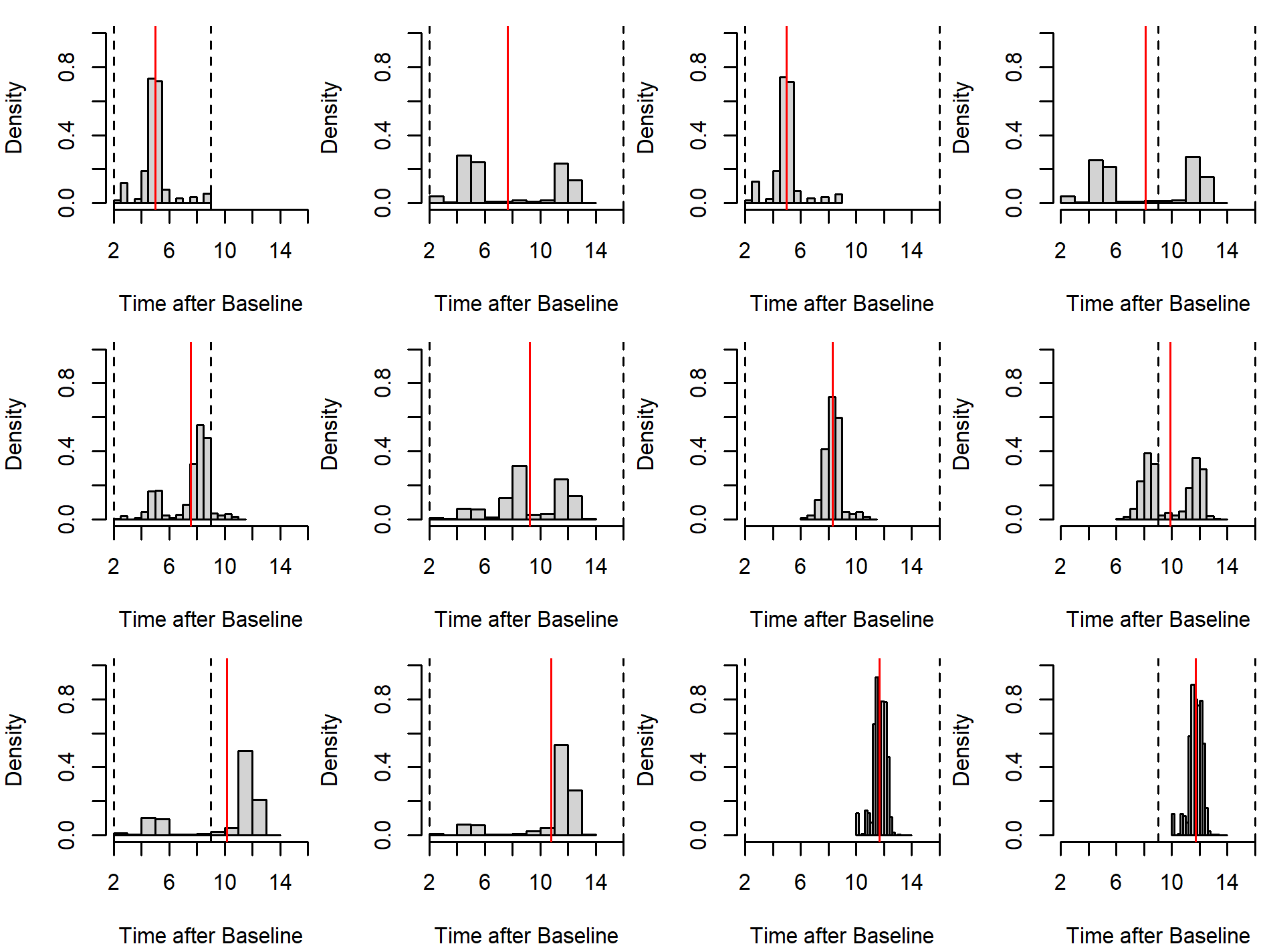

√

√

√

√

Late

Average

Early

Although we splited the data into 2-9 year and 9-16 follow-up groups according to the time interval between the subsequential cognitive tests and baseline assessment, 31,180 participants were presented in both groups. Therfore, selecting which measurment to use when analysing mtDNA-CN effects on cognitive decline across the full 16-year period can introduce statistical noise due to the varying time intervals. This figure displays the distribution of time interval for each subgroup, based on the combination of targeting follow-up period (2-9 years, 9-16 years, and 2-16 years) and the measurement used for those overlapping samples (early = measurement from 2-9 years, late = measurement from 9-16 years, and average = mean of both time points). Continuous referes to the ‘decline’ identified using three data points. Red line indicate the distribution mean. Dotted line shows the expected follow-up window. Ticks highlight the best solution. This figure shows that for those 31,180 overlapping samples, the appropriate measurements to use are: 2-9 year measurements, 9-year measurements, and average measurements for studying the mtDNA-CN effects on cognitive decline in 2-9 year, 9-16 year, and the full 2-16 year follow-up period, respectively. This ensures that, (a) no inclusion of data outside the target time window, (b) the mean of time interval aligns closely with the mean of time window, and (c) the distribution is more centered around the mean. The influence of using different measurements on estimation of mtDNA-CN effects is available in **Supplementary Table 5**.
